# Comparison of Laboratory-Based and Combined Clinical-Laboratory Models for Predicting Complicated Appendicitis: Incremental Value of the Prognostic Nutritional Index

**DOI:** 10.64898/2026.07.30.26359322

**Authors:** Daisuke Miura

**Author notes:** Corresponding author Daisuke Miura, PhD, Department of Ultrasound and Clinical Laboratory, Fukuoka Tokushukai Hospital 4-5 Sugu Kita, Kasuga, Fukuoka 816-0864, Japan.

## Abstract

**Background:** Early identification of complicated appendicitis is essential for appropriate management, yet accurate preoperative risk stratification remains challenging. Whether objective laboratory-based models can achieve predictive performance comparable to models incorporating clinical findings remains unclear.

**Objective:** To compare clinical, laboratory-based, and combined clinical-laboratory prediction models for complicated appendicitis and to evaluate the incremental value of the Prognostic Nutritional Index (PNI).

**Methods:** This retrospective study included 1,132 consecutive patients with acute appendicitis. Cases were classified as uncomplicated or complicated appendicitis based on pathological findings and imaging. Three multivariable logistic regression models were developed: a clinical model (Model A), a laboratory-based model (Model B), and a combined clinical-laboratory model (Model C). Model discrimination was assessed using the area under the receiver operating characteristic curve (AUC), and AUCs were compared using DeLong’s test.

**Results:** Complicated appendicitis was identified in 341 patients (30.1%). Higher C-reactive protein, higher neutrophil percentage, and lower PNI were independent predictors of complicated appendicitis. The AUCs were 0.791 for Model A, 0.847 for Model B, and 0.857 for Model C. Model B significantly outperformed Model A (*P* = 0.0020), whereas no significant difference was observed between Models B and C (*P* = 0.0816). PNI remained an independent predictor after adjustment for conventional inflammatory markers.

**Conclusion:** A laboratory-based prediction model incorporating PNI demonstrated discrimination comparable to that of a combined clinical-laboratory model while relying exclusively on routinely available objective laboratory parameters. These findings support laboratory-based risk stratification as a practical adjunct to clinical assessment for the early identification of complicated appendicitis.

**Article Summary:** *Why is this topic important?:* Early identification of complicated appendicitis is essential for timely surgical decision-making and appropriate resource utilization. However, the optimal balance between predictive performance and practical implementation of preoperative risk models remains uncertain.

*What does this study attempt to show?:* This study compared clinical, laboratory-based, and combined clinical-laboratory prediction models for complicated appendicitis and evaluated the incremental value of the Prognostic Nutritional Index (PNI). It also assessed whether an objective laboratory-based model could achieve predictive performance comparable to that of a combined clinical-laboratory model.

*What are the key findings?:*

- A laboratory-based model incorporating PNI demonstrated discrimination comparable to that of the combined clinical-laboratory model.
- PNI remained an independent predictor after adjustment for conventional inflammatory markers.
- The combined model achieved the best overall predictive performance, whereas the laboratory-based model offered greater simplicity and potential for standardized implementation.

*How is patient care impacted?:*

- Objective laboratory-based risk stratification may support early identification of patients at high risk for complicated appendicitis.
- A laboratory-based model may be particularly useful in settings where advanced imaging is limited or ultrasonography expertise is unavailable.
- Routine laboratory data may facilitate standardized preoperative risk assessment and clinical decision-making.

## Introduction

Acute appendicitis is one of the most common causes of acute abdominal pain requiring emergency evaluation and surgical intervention (1,2). Early identification of complicated appendicitis is essential because it guides treatment decisions, including the need for urgent surgery and perioperative management, and may reduce the risk of postoperative complications (3–5). Although contrast-enhanced computed tomography (CT) is widely regarded as the reference standard for diagnosing acute appendicitis, its routine use may be limited by radiation exposure, cost, and limited availability, whereas the diagnostic performance of ultrasonography (US) remains highly operator dependent (6,7). Therefore, there is a continuing need for simple, objective, and readily available prediction tools to support the early risk stratification of complicated appendicitis.

Several clinical scoring systems, including the Alvarado score and the Appendicitis Inflammatory Response score, have been developed to estimate the likelihood of acute appendicitis using combinations of clinical findings and laboratory parameters (8–10). In addition to clinical scoring systems, US and CT are widely used to establish the diagnosis and evaluate disease severity (10). However, accurate preoperative differentiation between uncomplicated and complicated appendicitis remains challenging.

Patients with complicated appendicitis have higher morbidity, longer hospital stays, and are more likely to require urgent surgery or other invasive interventions (4,11). Consequently, laboratory markers of systemic inflammation, such as the white blood cell count (WBC) and C-reactive protein (CRP), are widely incorporated into predictive models (8–10). Composite inflammatory indices derived from routine blood tests, such as the neutrophil-to-lymphocyte ratio (NLR) and platelet-to-lymphocyte ratio (PLR), have also been investigated as predictors of disease severity (12,13).

However, these biomarkers primarily reflect the magnitude of the inflammatory response and may be influenced by the timing of disease presentation and interindividual variability. Consequently, their ability to reliably discriminate between uncomplicated and complicated appendicitis remains limited, underscoring the need for additional biomarkers that capture broader aspects of the host response.

Despite these advances, current approaches remain imperfect for preoperative risk stratification of complicated appendicitis. Composite indices derived from routine laboratory data have attracted attention as potential predictive biomarkers. Commonly used inflammatory indices, such as the NLR and PLR, primarily reflect inflammatory activity and have shown inconsistent predictive performance across studies (12,13). In contrast, the Prognostic Nutritional Index (PNI), which incorporates serum albumin concentration and peripheral lymphocyte count, may provide complementary information by reflecting host immune-nutritional status (14). Although PNI has been widely investigated in oncology and chronic inflammatory diseases (15,16), its role in acute abdominal emergencies, particularly in distinguishing complicated from uncomplicated appendicitis, remains poorly defined.

Although recent studies have explored PNI as a predictor of complicated appendicitis, these investigations have generally been limited by relatively small sample sizes or heterogeneous diagnostic criteria, restricting the generalizability of their findings (17,18). Therefore, whether laboratory-based models incorporating PNI can achieve predictive performance comparable to that of combined clinical-laboratory models for preoperative risk stratification of complicated appendicitis in a large, well-defined cohort remains unknown.

Therefore, the purpose of this study was to develop and compare clinical, laboratory-based, and combined clinical-laboratory prediction models for complicated appendicitis in a large, well-defined cohort of patients with acute appendicitis. We further hypothesized that incorporating PNI, as a marker of host immune-nutritional status, would provide independent and incremental predictive value beyond conventional inflammatory markers. Model performance was comprehensively evaluated to determine whether a laboratory-based model incorporating PNI could achieve predictive performance comparable to that of a combined clinical-laboratory model.

## Methods

### Study Design and Participant Recruitment

This retrospective observational study was conducted at Fukuoka Tokushukai Hospital (Fukuoka, Japan). Consecutive patients who presented between April 2017 and June 2025 with suspected acute appendicitis, based on clinical assessment and US and/or CT findings, were screened for eligibility.

A total of 1,191 patients were screened. Of these, 50 were excluded because an alternative diagnosis was established during the diagnostic workup, and 9 were excluded because laboratory data at initial presentation were unavailable. Consequently, 1,132 patients with acute appendicitis were included in the final analysis (**Figure 1**).

**Figure 1.**
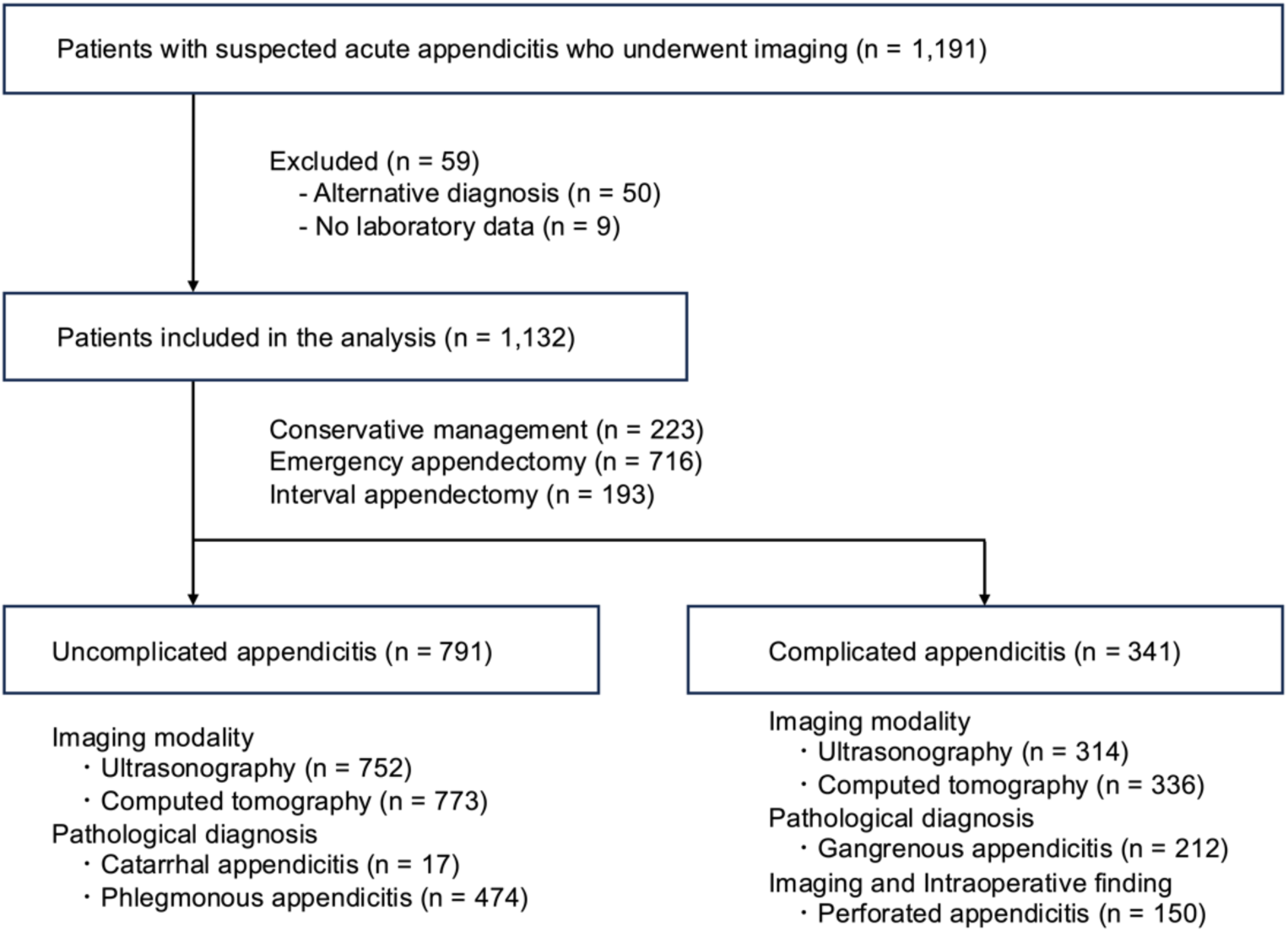
Flowchart of Patient Selection and Study Cohort. A total of 1,191 patients with suspected acute appendicitis were screened. After exclusion of patients with an alternative diagnosis and those without laboratory testing at initial presentation, 1,132 patients were included in the final analysis. **Abbreviations:** CT, computed tomography; US, ultrasonography.

The study was approved by the Institutional Review Board of Fukuoka Tokushukai Hospital (Approval No. 260301), which waived the requirement for informed consent because of the retrospective study design.

### Definition of Uncomplicated and Complicated Appendicitis

Uncomplicated appendicitis was defined by the following criteria: (1) preservation of the normal layered structure of the appendiceal wall on US; (2) absence of perforation on intraoperative findings when surgery was performed; and (3) pathological diagnosis of catarrhal or phlegmonous appendicitis.

Complicated appendicitis was defined as gangrenous or perforated appendicitis. Gangrenous appendicitis was diagnosed by pathological examination. Perforated appendicitis was diagnosed when perforation was identified by intraoperative findings or imaging studies, including US and/or contrast-enhanced CT.

In surgically treated patients, disease classification was based on pathological examination together with intraoperative findings. In patients managed nonoperatively, classification was based on imaging findings and the clinical course. Imaging findings were used exclusively for outcome classification and were not included as explanatory variables in any predictive model.

### Clinical and Laboratory Variables

Clinical variables collected at initial presentation included age, sex, body temperature, time from symptom onset to hospital arrival, gastrointestinal symptoms (nausea, vomiting, and diarrhea), and physical examination findings (McBurney’s point tenderness, muscular defense, and rebound tenderness). Fever was defined as a body temperature ≥37.3°C.

Laboratory variables collected at initial presentation included CRP, WBC, neutrophil percentage, lymphocyte percentage, platelet count, PLR, total bilirubin, and PNI. PNI was calculated as 10 × serum albumin (g/dL) + 0.005 × peripheral lymphocyte count (/mm³).

### Model Construction

Three multivariable logistic regression models were constructed to predict complicated appendicitis using different combinations of clinical and laboratory variables. Model A (clinical model) included clinical variables only, Model B (laboratory model) included laboratory variables only, and Model C (combined clinical-laboratory model) incorporated both clinical and laboratory variables. Candidate variables identified in the univariable analyses were considered for inclusion in the multivariable models based on statistical significance and clinical relevance. Final variable selection was based on statistical significance and assessment of multicollinearity using the variance inflation factor (VIF).

### Statistical Analysis

Continuous variables were summarized as medians with interquartile ranges (IQRs). Normality was assessed using the Shapiro–Wilk test. Because not all continuous variables followed a normal distribution, nonparametric statistical methods were consistently applied throughout the analyses. Categorical variables were expressed as frequencies and percentages. Between-group comparisons were performed using the Mann–Whitney U test for continuous variables and the chi-square test or Fisher’s exact test for categorical variables, as appropriate.

Multivariable logistic regression analysis was performed to identify independent predictors of complicated appendicitis. Because five patients had incomplete data for variables included in the multivariable models, these analyses were performed using complete-case data. Adjusted odds ratios (ORs) with 95% confidence intervals (CIs) were calculated. Model discrimination was evaluated using the area under the receiver operating characteristic curve (AUC), and comparisons between AUCs were performed using the DeLong test. Model calibration was assessed using calibration plots, and overall prediction accuracy was evaluated using the Brier score. Clinical utility was evaluated using decision curve analysis. Model fit was additionally assessed using the Akaike information criterion (AIC).

All statistical analyses were performed using StatFlex (version 7.0; Artech, Osaka, Japan). Internal validation was performed using bootstrap resampling with 1,000 iterations to estimate optimism-corrected model performance. Calibration plots, decision curve analysis, and internal validation were additionally performed using Python (Python Software Foundation, Wilmington, DE, USA). A two-sided *P* value < 0.05 was considered statistically significant.

## Results

### Patient Characteristics

A total of 1,132 patients with acute appendicitis were included in the analysis, of whom 341 (30.1%) were classified as having complicated appendicitis.

US was performed in 1,075 patients (95.0%), and CT, with or without intravenous contrast enhancement, was performed in 1,100 patients (97.2%).

Among patients who underwent emergency appendectomy, pathological examination demonstrated catarrhal appendicitis in 17 patients and phlegmonous appendicitis in 474 patients, both classified as uncomplicated appendicitis. Gangrenous appendicitis was identified in 212 patients, classified as complicated appendicitis. In addition, perforated appendicitis was diagnosed in 150 patients based on imaging findings and/or intraoperative macroscopic findings. Baseline demographic, clinical, and laboratory characteristics are summarized in **Table 1**.

**Table 1.**
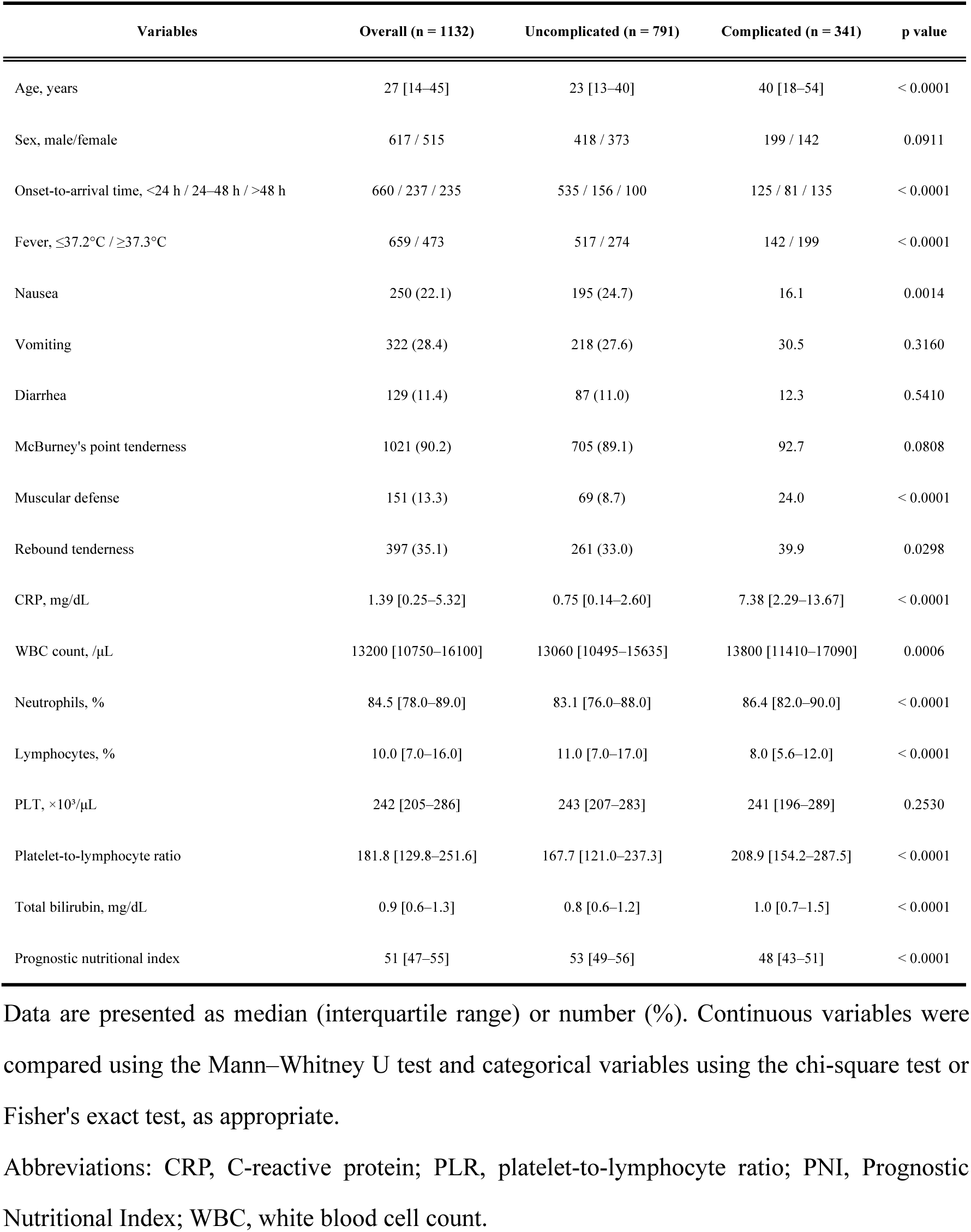
Baseline Clinical and Laboratory Characteristics of Patients With Uncomplicated and Complicated Appendicitis.

### Univariable Analysis of Clinical and Laboratory Variables

Clinical and laboratory variables were compared between patients with uncomplicated and complicated appendicitis using univariate analyses (**Table 1**).

Among clinical variables, patients with complicated appendicitis were significantly older and had a longer onset-to-arrival time than those with uncomplicated appendicitis (both *P* < 0.0001). Fever (≥37.3°C), muscular defense, and rebound tenderness were significantly more frequent in the complicated group (*P* < 0.0001, *P* < 0.0001, and *P* = 0.0298, respectively), whereas nausea was significantly less frequent (*P* = 0.0014). Sex, vomiting, diarrhea, and McBurney’s point tenderness did not differ significantly between the two groups.

Among laboratory variables, patients with complicated appendicitis had significantly higher CRP, WBC, neutrophil percentage, PLR, and total bilirubin, whereas lymphocyte percentage and PNI were significantly lower than in patients with uncomplicated appendicitis (all *P* < 0.0001, except WBC count, *P* = 0.0006). Platelet count did not differ significantly between the two groups.

### Multivariable Logistic Regression Analysis for the Identification of Independent Predictors of Complicated Appendicitis

Multivariable analyses were based on 1,127 patients with complete data. **Table 2** presents the results of the multivariable logistic regression analyses for identifying independent predictors of complicated appendicitis. Three models were constructed: a clinical model (Model A), a laboratory model (Model B), and a combined clinical–laboratory model (Model C).

**Table 2.**
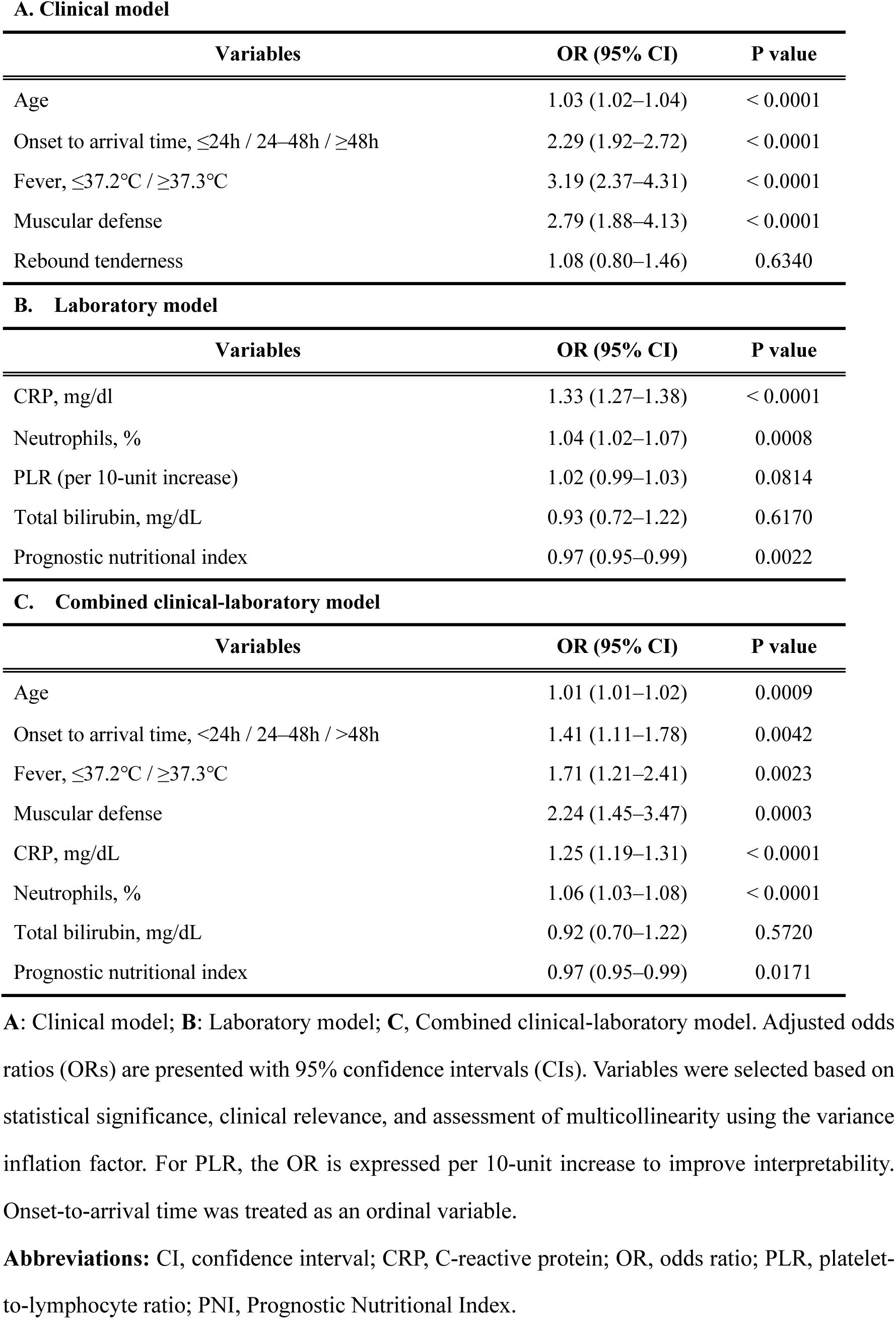
Multivariable Logistic Regression Models for Predicting Complicated Appendicitis.

**A. Clinical model**
| Variables | OR (95% CI) | P value |
| --- | --- | --- |
| Age | 1.03 (1.02–1.04) | < 0.0001 |
| Onset to arrival time, ≤24h / 24–48h / ≥48h | 2.29 (1.92–2.72) | < 0.0001 |
| Fever, ≤37.2°C / ≥37.3°C | 3.19 (2.37–4.31) | < 0.0001 |
| Muscular defense | 2.79 (1.88–4.13) | < 0.0001 |
| Rebound tenderness | 1.08 (0.80–1.46) | 0.6340 |

**B. Laboratory model**
| Variables | OR (95% CI) | P value |
| --- | --- | --- |
| CRP, mg/dl | 1.33 (1.27–1.38) | < 0.0001 |
| Neutrophils, % | 1.04 (1.02–1.07) | 0.0008 |
| PLR (per 10-unit increase) | 1.02 (0.99–1.03) | 0.0814 |
| Total bilirubin, mg/dL | 0.93 (0.72–1.22) | 0.6170 |
| Prognostic nutritional index | 0.97 (0.95–0.99) | 0.0022 |

**C. Combined clinical-laboratory model**
| Variables | OR (95% CI) | P value |
| --- | --- | --- |
| Age | 1.01 (1.01–1.02) | 0.0009 |
| Onset to arrival time, <24h / 24–48h / >48h | 1.41 (1.11–1.78) | 0.0042 |
| Fever, ≤37.2°C / ≥37.3°C | 1.71 (1.21–2.41) | 0.0023 |
| Muscular defense | 2.24 (1.45–3.47) | 0.0003 |
| CRP, mg/dL | 1.25 (1.19–1.31) | < 0.0001 |
| Neutrophils, % | 1.06 (1.03–1.08) | < 0.0001 |
| Total bilirubin, mg/dL | 0.92 (0.70–1.22) | 0.5720 |
| Prognostic nutritional index | 0.97 (0.95–0.99) | 0.0171 |

**A:** Clinical model; **B:** Laboratory model; **C,** Combined clinical-laboratory model. Adjusted odds ratios (ORs) are presented with 95% confidence intervals (CIs). Variables were selected based on statistical significance, clinical relevance, and assessment of multicollinearity using the variance inflation factor. For PLR, the OR is expressed per 10-unit increase to improve interpretability. Onset-to-arrival time was treated as an ordinal variable.
**Abbreviations:** CI, confidence interval; CRP, C-reactive protein; OR, odds ratio; PLR, platelet- to-lymphocyte ratio; PNI, Prognostic Nutritional Index.

Model A: Older age, longer onset-to-arrival time, presence of fever, and muscular defense were independently associated with complicated appendicitis, whereas rebound tenderness was not.

Model B: Higher CRP, higher neutrophil percentage, and lower PNI were independently associated with complicated appendicitis. Lower PNI remained an independent predictor after adjustment for other laboratory variables (OR 0.97, 95% CI 0.95–0.99, *P* = 0.0022). PLR and total bilirubin were not independently associated with complicated appendicitis.

Model C: Older age, longer onset-to-arrival time, presence of fever, muscular defense, higher CRP, higher neutrophil percentage, and lower PNI remained independently associated with complicated appendicitis. Lower PNI remained an independent predictor after adjustment for the other variables in the combined model (OR 0.97, 95% CI 0.95–0.99, *P* = 0.0171).

### Model Performance

**Figure 2** shows the ROC curves for the three prediction models. The AUC was 0.791 for Model A, 0.847 for Model B, and 0.857 for Model C, demonstrating progressively improved discrimination across the three models.

**Figure 2.**
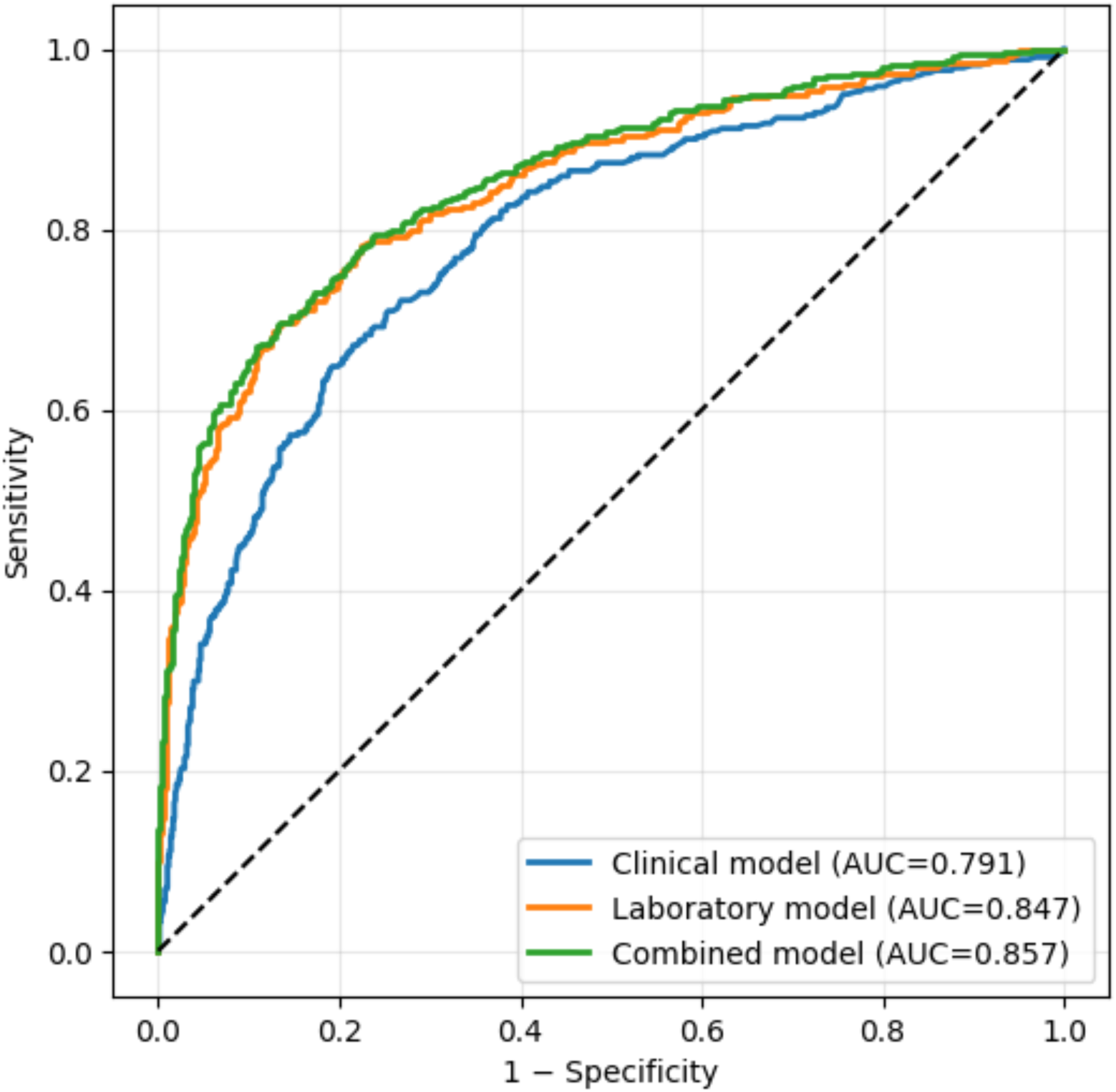
Receiver Operating Characteristic Curves for the Three Prediction Models. Receiver operating characteristic (ROC) curves comparing the discriminative performance of the clinical (Model A), laboratory (Model B), and combined clinical-laboratory (Model C) prediction models. Pairwise comparisons of AUCs were performed using DeLong’s test.

Pairwise comparisons of the AUCs were performed using DeLong’s test. Model B demonstrated a significantly higher AUC than Model A (*P* = 0.0020), and Model C also showed a significantly higher AUC than Model A (*P* < 0.0001). No significant difference in AUC was observed between Models B and C (*P* = 0.0816).

Internal validation using 1,000 bootstrap resamples demonstrated minimal optimism, indicating little evidence of model overfitting. The optimism-corrected AUCs were 0.791 for the clinical model, 0.846 for the laboratory model, and 0.853 for the combined model.

Model fit was additionally assessed using AIC. Model C demonstrated the lowest AIC (930.6), followed by Model B (958.9) and Model A (1125.6), indicating the best overall model fit.

**Figure 3** shows the calibration plots for the three prediction models. All models demonstrated good agreement between predicted and observed probabilities across the range of predicted risks. Models B and C demonstrated good agreement with the ideal calibration line, whereas Model C showed the closest agreement overall. Model A exhibited slightly greater deviation from the ideal line, particularly at higher predicted probabilities.

**Figure 3.**
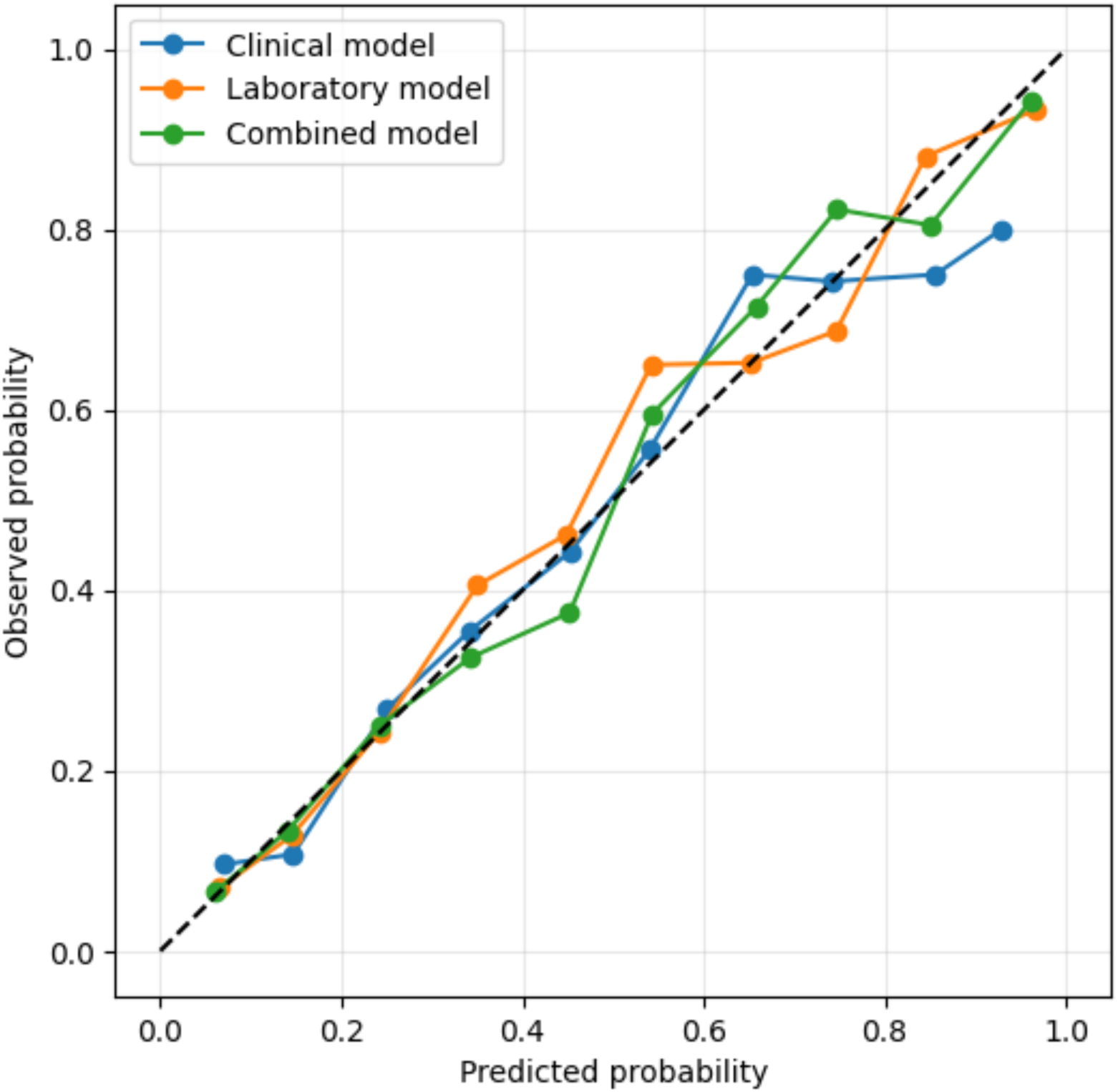
Calibration Plots for the Three Prediction Models. Calibration plots illustrating the agreement between predicted and observed probabilities of complicated appendicitis for the clinical (Model A), laboratory (Model B), and combined clinical-laboratory (Model C) prediction models. The dashed diagonal line represents perfect calibration.

Overall prediction accuracy was further assessed using the Brier score. Model C achieved the lowest Brier score (0.1265), followed by Model B (0.1313) and Model A (0.1603), indicating progressively improved overall prediction accuracy across the three models.

The decision curve analysis is presented in **Figure 4**. Across clinically relevant threshold probabilities, Models B and C consistently provided higher net benefit than Model A. Model C achieved the greatest net benefit over most threshold probabilities, whereas the difference between Models B and C was modest. All three models demonstrated greater clinical utility than the “treat-all” and “treat-none” strategies across a broad range of threshold probabilities.

**Figure 4.**
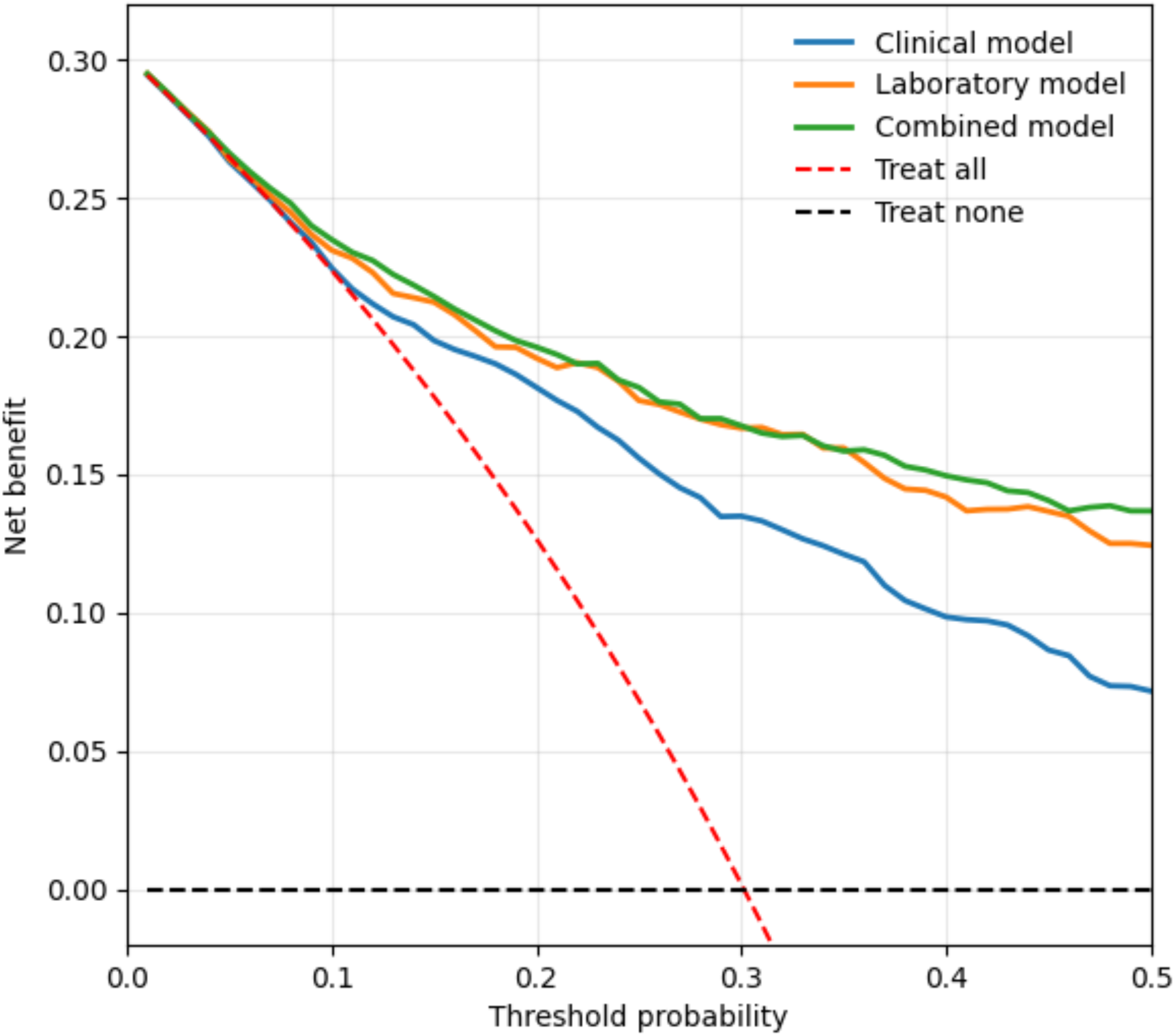
Decision Curve Analysis of the Three Prediction Models. Decision curve analysis comparing the clinical utility of the clinical (Model A), laboratory (Model B), and combined clinical-laboratory (Model C) prediction models across a range of threshold probabilities. The red line represents the treat-all strategy, and the horizontal black line represents the treat-none strategy.

## Discussion

### Principal Findings

In this large cohort study, the laboratory-based model (Model B) demonstrated strong discriminatory ability for predicting complicated appendicitis, comparable to that of the combined clinical–laboratory model (Model C). Although Model C consistently showed the best overall performance, including discrimination, model fit, calibration, overall prediction accuracy, and clinical utility, its discriminative advantage over Model B was not statistically significant. Notably, PNI remained an independent predictor of complicated appendicitis after adjustment for conventional inflammatory markers, highlighting the potential contribution of host-related factors beyond the inflammatory response alone.

### Interpretation of Key Predictors

Among the laboratory parameters, CRP emerged as the most influential predictor of complicated appendicitis, likely reflecting the overall magnitude of the inflammatory response and its temporal accumulation. An elevated neutrophil percentage may represent progression of bacterial infection and activation of the innate immune response.

In contrast, PNI may capture a distinct aspect of disease severity that is not fully explained by conventional inflammatory markers. Although PNI has traditionally been interpreted as a marker of nutritional status, its predictive value in the present study is more plausibly attributable to lymphocyte depletion induced by acute systemic inflammatory stress rather than hypoalbuminemia alone. Given the acute nature of appendicitis, substantial reductions in serum albumin are unlikely to occur over a short time course, whereas lymphocyte counts are known to decrease rapidly in response to stress, inflammation, and immune dysregulation.

Importantly, PNI remained an independent predictor complicated appendicitis after adjustment for CRP and neutrophil percentage, suggesting that it may reflect host-related vulnerability beyond the intensity of inflammation itself, potentially related to immune suppression or stress-induced immunological exhaustion. This independence implies that PNI may serve as a surrogate marker of impaired host response, thereby identifying patients in whom acute inflammation has already exerted systemic effects.

In addition to laboratory markers, a longer onset-to-arrival time was independently associated with complicated appendicitis, supporting the concept that delayed presentation facilitates progression from localized to advanced disease. Taken together, these findings suggest that parameters reflecting inflammatory burden, host immune resilience, and disease chronology provide complementary information that may improve risk stratification for complicated appendicitis.

### Comparison of Prediction Models

An important finding of this study was that Model B demonstrated discriminatory performance comparable to that of Model C, as confirmed by formal AUC comparisons. Although the addition of clinical variables resulted in a numerically higher AUC, this improvement did not reach statistical significance. These findings suggest that objective laboratory parameters capture much of the information required to identify complicated appendicitis, likely reflecting the underlying inflammatory burden and disease severity in a more standardized manner than clinical assessment alone.

Nevertheless, clinical variables such as fever and muscular defense remained independently associated with complicated appendicitis in Model C, indicating that clinical evaluation continues to provide complementary information, particularly in characterizing disease presentation. In addition to demonstrating the highest discrimination, Model C also showed the best model fit, calibration, overall prediction accuracy, and clinical utility. However, the improvements over Model B were generally modest, particularly with respect to discrimination, for which no statistically significant difference was observed.

Taken together, these findings highlight the central role of objective laboratory data in early risk stratification while supporting the continued importance of clinical assessment as a complementary adjunct that may further improve model performance and clinical decision-making.

Although Model C consistently achieved the best overall performance, the modest incremental benefit over Model B should be interpreted in the context of model simplicity and implementability. Because Model B relies exclusively on routinely available objective laboratory parameters, it may be more readily standardized, automated, and incorporated into routine clinical workflows or electronic decision support systems.

### Clinical Implications

From a clinical perspective, although Model C demonstrated the best overall predictive performance, Model B maintained strong predictive performance using objective laboratory parameters alone. Decision curve analysis further suggested that both models may provide clinical benefit across a broad range of threshold probabilities, particularly when determining the need for urgent imaging or surgical consultation. These findings indicate that risk stratification may be feasible using routinely available objective laboratory data alone.

These findings are particularly relevant in settings with limited access to advanced imaging, such as small or resource-limited clinics, where computed tomography may not be readily available and ultrasonography is highly operator-dependent. In such settings, laboratory-based risk stratification may provide an objective and readily available adjunct to clinical assessment, thereby helping to identify patients who warrant urgent referral or further imaging evaluation.

Because Model B relies exclusively on routinely available laboratory parameters, it also has the potential to be readily automated within electronic health record systems or clinical decision support tools. Automated calculation of individualized risk immediately after laboratory results become available may facilitate standardized early risk stratification while minimizing interobserver variability in clinical assessment, thereby supporting timely clinical decision-making in emergency care.

### Limitations

Several limitations should be acknowledged. First, this was a retrospective single-center study, which may limit the generalizability of the findings. Internal validation using bootstrap resampling demonstrated minimal optimism, supporting the robustness of the developed models. However, external validation remains necessary before clinical implementation.

Second, some degree of incorporation bias cannot be excluded because clinical findings and laboratory results inevitably influenced diagnostic and treatment decisions. However, this reflects routine emergency practice and therefore enhances the clinical relevance of the study.

Finally, disease classification in patients managed non-operatively relied on imaging findings and clinical course rather than pathological confirmation, which may have resulted in some misclassification. Future studies should focus on external validation in independent cohorts to confirm the generalizability, and clinical applicability of these prediction models.

## Conclusions

In conclusion, a laboratory-based prediction model incorporating PNI demonstrated discriminatory performance comparable to that of a combined clinical-laboratory model for identifying complicated appendicitis. Although the combined model achieved the best overall performance, the laboratory-based model offers important practical advantages because it relies exclusively on routinely available objective laboratory parameters. These findings support the use of laboratory-based risk stratification as a practical adjunct to clinical assessment and provide a foundation for future standardized or automated clinical decision support in emergency practice.

## Acknowledgements

None.

## Ethics Approval

This retrospective study was approved by the Institutional Review Board of Fukuoka Tokushukai Hospital (Approval No. 260301) and was conducted in accordance with the Declaration of Helsinki.

## Informed Consent

The requirement for informed consent was waived by the Institutional Review Board because of the retrospective nature of the study.

## Data Availability

The datasets generated and/or analyzed during the current study are available from the corresponding author on reasonable request.

## Author Contributions

Daisuke Miura: Conceptualization, Methodology, Data curation, Formal analysis, Visualization, Writing – original draft.

